# OIICS coding of agricultural injuries mined from Maine PCR records (2008-2022) reveals significant differences in injury source, event, and nature by age group and sex

**DOI:** 10.1101/2025.09.24.25336586

**Authors:** Laura E. Jones, Megan Kern, Cristina S. Hansen-Ruiz, Nicole Krupa, Paul Jenkins, Erika Scott

## Abstract

**Objectives:** Agricultural injuries are known to be under-reported in existing surveillance systems. OIICS codes are a standardized classification system developed by the Bureau of Labor Statistics (BLS) which ensure consistency in reporting and analysis of workplace incidents over time across industry sectors. Our study examines OIICS coded injuries obtained via mining emergency response (Pre-Hospital Care Report) records (PCRs) to improve tracking, documentation, and understanding of agricultural injury trends.

**Methods:** We analyzed frequencies of OIICS subcodes for Primary Injury Source, Event/Exposure, Nature of Injury, and Body Part classifications for 1,583 injuries among agricultural workers in Maine, spanning January 2008 to December 2022. To streamline the dataset and subsequent analysis, subcodes within each category were thematically grouped. We summarized and visualized grouped code frequencies by subject sex, age category, season of injury, and study subperiod. Chi-square tests were used to assess differences in injury patterns by sex and age group.

**Results:** Reported injuries increased over time from 420 in 2008–2011 to 631 in 2019–2022. The most frequently reported classifications were: ‘Tractors/PTOs’ (Injury Source), ‘Fall’ (Event), ‘Multiple parts’ (Body Part), and ‘Pain’ (Nature of Injury). A marked increase in ‘Nonclassifiable’ Source subcodes and ‘Fall’ Event subcodes was observed in 2019–2022 relative to earlier periods. Significant differences by sex were found for injury Event subcodes: The most frequent source of injuries for females were animals, versus objects and equipment being the most frequent source for males. Nature of Injury also varied significantly by sex. All four OIICS categories (Source, Event, Nature, Body Part) showed significant variation by age group. Older subjects reported more injuries due to falls and overexertion, while younger were more frequently subject to exposure, intentional self-injury, injury in fires, and injuries involving farm vehicles and equipment.

**Conclusion:** Injury counts rose across each successive study period. All injury subcodes differed significantly by age category, while injury Event and Body Part codes also varied significantly by sex. This suggests that injury risks are not uniform across demographics, and tailored safety interventions by sex and age group may be more effective in reducing agricultural injuries.

## Introduction

Agricultural injuries are under-reported in many occupational health surveillance systems that maintain work-related morbidity databases.^1,2^ Smaller farms are not subject to the same oversight and reporting from the Occupational Safety and Health Administration (OSHA) as are farms with greater than 10 employees.^3^ Therefore, critical data from a large swath of agricultural operations are absent from federal occupational injury reporting. Given that the over ninety percent (94.8%) of farms in the state of Maine have fewer than 10 employees, alternative methods to capture otherwise unreported agricultural injuries are needed.^4^ This is especially vital given that fatal occupational injury data demonstrates that agriculture remains one of the top ten most dangerous civilian occupations in the United States.^5^

To fill the gaps in reporting, injury and occupational epidemiologists have employed a variety of methods including scraping the internet,^6-8^ utilizing surveys^9-11^ and interviews,^12^ as well as scouring insurance^13,14^ and administrative health records.^15-18^ In order for these disparate data sources to be comparable and compatible, a common coding scheme was devised specifically for occupational injuries and illnesses.^19^ The Occupational Injury and Illnesses Classification System (OIICS) was developed by the Bureau of Labor Statistics and is used by occupational injury researchers and government statisticians to uniformly describe the various aspects of the injury scenario.^20-22^

One promising data source for agricultural injuries is the pre-hospital care report, or PCR. This report, generated by emergency medical technicians and paramedics following an ambulance response, contains a detailed free-text narrative that describes, in the responders own language, what they encountered at the scene, details of the patient’s conditions, treatment, and much more.^23-25^ Since responders arrive at the scene of the injury event, they often have firsthand information about what caused the incident, which is essential information for the injury prevention practitioner. The purpose of this study was to examine OIICS coded injuries obtained via mining emergency response (Pre-Hospital Care Report) records (PCRs) to improve tracking, documentation, and understanding of agricultural injury trends.

## Methods

### Data

We employ a dataset derived from processed and classified PCR records from the state of Maine for years 2008 through 2022. Cleaning and processing is described in detail by Hirabayashi and others^26^, and the machine learning workflow used to label it is described by Scott and others^27^. We were unable to acquire records for 2017, and those from 2018 were omitted due to coding conflicts. Our dataset comprises coded records labeled by our machine learning workflow as follows: *confirmed acute or traumatic agricultural injuries, confirmed acute or traumatic, suspected agricultural*, and *suspected acute or traumatic, confirmed agricultural injuries*. After labeling, the records were sent to a team of human coders who determined full Occupational Injury and Illness Classification (OIICS) codings (v. 2.01) for each record. From these, we examine frequencies of OIICS Primary Injury Source, Event/Exposure, Injury Nature and Body Part codings for 1,583 injuries occurring among agricultural workers in the state of Maine from January 2008 to December 2022. We omit 21 coded injuries occurring between January and April of 2023 as this is an incomplete year. Missing units for the remaining sample are approximately 2.7%, and affect age determinations.

### Covariates

Our dataset includes sex (Male, Female, Unknown), age (years), injury month and year (2008 – 2016, 2019 – 2022), and four coded OIICS levels that describe the injury and its circumstances. Study population subjects were categorized by age as follows: age no more than 20 years, age 21 to 40 years, age 41 to 60 years, age 61 to 80 years, and age 81 and above. For one visualization task we combine the last two age categories due to low counts in the fifth age category (81 and above). Due to missing/unavailable data described above for years 2017 and 2018, we analyze our dataset in three roughly equal periods bracketing the break: 2008-2011; 2012-2016, and 2019-2022.

### OIICS codings

OIICS codes are a standardized classification system used to describe the characteristics of work-related injuries, illnesses, and fatalities.^19^ Developed by the Bureau of Labor Statistics (BLS), these codes are used by BLS data programs such as the Survey of Occupational Injuries and Illnesses (SOII) and Census of Fatal Occupational Injuries (CFOI) but also by occupational epidemiologists and researchers. They help ensure consistency in reporting and analyzing workplace incidents across industries and over time.

OIICS classification is divided into four major structures, each with hierarchical subcodes:

1. **Primary and Secondary Source of Injury or Illness** – Indicates the object, substance, or person that directly caused the injury (e.g., machinery, chemicals). We limit our study to primary sources.
2. **Event or Exposure** – Describes how the injury occurred (e.g., fall, overexertion, contact with object).
3. **Nature of Injury or Illness** – Describes the physical characteristics (e.g., fracture, burn, sprain).
4. **Part of Body Affected** – Identifies the body part involved (e.g., head, trunk, legs).

Each structure uses a multi-digit code, where each digit or group of digits adds specificity. For example, a 4-digit code might start broad with the first digits and become more detailed with each level (i.e., additional digits). Comprehensive OIICS coding analysis includes describing an injury scenario using one code from each of the four structures listed above. This coding structure provides a complete picture of the incident, including what happened, what body part(s) were injured, the type of injury sustained, and what caused it. However, in many analyses, only selected codes are used—often due to relevance to the research question, data availability or quality, or simplification for statistical modeling or reporting. The reliability of the OIICS coding conducted by this research team has been previously established.^20^

### Code groupings

We grouped codes thematically under each of the four structures above to reduce dimension of the dataset and facilitate analysis. For primary injury **source**, themes included tractors, farm vehicles and their components (parts), machinery, work surfaces such as ramps, floors and stairs, structures, large animals such as horses or cattle, temperature extremes; for injury **event**, themes included falls, being struck by equipment or animals, overexertion, roadway and non-roadway vehicular incidents, contact or compression by an object or equipment, temperature extremes, intentional injury. Thematic categories included in the labeled **nature** of injury group included pain, single or multiple traumatic injury; cuts, open wounds, bruises or fractures; chest pain, poisoning, and temperature extremes; body **part** injured label category included separate upper and lower body themes, the trunk excluding chest, the chest, legs, head, a category for multiple body parts and a generic ‘body’ category. Miscellaneous (‘misc.’) categories were included for items that did not fit under any given label and for which there were too few instances to justify another thematic label. See Supplemental Table 1 for a listing of numeric OIICS codes falling under each structure and thematic label.

### Visualizations

As with prior analysis, we define three roughly four-year periods for separate analysis due to the gap in the data comprising years 2017 and 2018: 2008 to 2011, 2012 to 2016 and 2019 to 2022. The second period (2012 to 2016) includes a fifth full year; the other two periods include four full years. We also omit injury counts from the partial year 2023, as including an incomplete year in the data processing introduced missing units and rendered it difficult to compare by period and season. We followed this strategy with a prior effort on the same dataset (Jones, Scott et al. 2025, submitted) as seasonal time series data require complete cycles for analysis.

We created visualizations of grouped codes across the three periods of interest (2008-2011, 2012-2016 and 2019-2022), and by season (Winter, Spring, Summer, Fall) to reveal notable changes in code frequencies by period and to visualize seasonal differences. Injury rates were found to differ strongly by age category (Jones, Scott et al. 2025b), and preliminary clustering analysis by multiple correspondence analysis (MCA) also showed strong differences in code distributions by subject sex. We thus created visualizations by sex. Visualizations for labeled primary source (Source1) and injury event (Event) themes are shown and discussed in the main text and those for injury nature (Nature) and body part (Part) are shown in Supplementary Information and discussed where relevant in the main text.

### Summary tables and bivariate analysis

Based on results from prior work (Jones, Scott et al. 2025b, submitted), we summarize participants by sex, age category and period, and summarize OIICS subcode distributions by sex (omits 29 subjects of unknown sex). We determined if differences in injury code distributions are significant by sex, age category, season and period using chi-squared tests. P-values for the chi-squared tests are determined via Monte Carlo with 10,000 simulations.

### Software

Data cleaning and wrangling were performed in the R programming language (R4.4.1) using tidyverse. We created visualizations using ggplot2 and pheatmaps in R.

## Results

### Code groupings

We grouped codes thematically to reduce dimension of the dataset and facilitate analysis. Please see supplementary Table S1 for a listing of codes grouped under each OIICS classification level (Source1, Event, Nature, Part). Our grouping process gathered 270 primary source codes under 15 thematic labels; 249 injury event codes under 17 thematic labels; 175 injury nature codes under 17 thematic labels, and 123 body part injured codes gathered under 11 labels. All reduced code levels included as part of the counts above a miscellaneous category for codes that appear at less than 5% frequency, and a ‘missing’ category for levels explicitly coded as ‘missing.’

### Heat map visualizations

Our heat maps are bivariate visualizations of cross-tabulations between a given grouped code level and a feature of interest such as study period, season, sex, or age category; essentially matrices of values presented as colored maps. Before performing cross-tabulations, code groupings are arranged from top (maximum) to bottom (minimum) in terms of total counts. Count maps are scaled to minimum (yellow) and maximum (purple) cell counts. In addition, in order to better understand the distributions of injury source, event, nature and body part injured by age, we have created *scaled* maps for age category. Here, we mean-standardize counts for each grouped code by sex for maps by sex. For age category, period and season, we scale by row to reveal which age category, period or season has the highest hit count for a particular code. The color key for the maps runs from yellow for low counts (below row mean for scaled maps) to purple for high counts (above row mean for scaled maps). Zero counts in unscaled maps show no color.

### Injury by sex

Some of the largest differences in primary injury source and event were between sexes as shown in the heat maps Figure 1. Left hand panels show count heat maps to demonstrate global peaks in primary source and event; right hand panels are scaled by sex to highlight the sources and events experienced most often by each sex. Men were more likely to show tractors/PTO’s as greatest primary injury source (Source1) whereas women were more likely to show ‘horses’ or ‘cattle’ as primary injury source (Figure 1). Both sexes showed ‘Fall’ as dominant event category, but males were then more likely to be struck by equipment while female subjects, struck by animals. Though the visualization suggests proportionally more head injuries in women, relative differences by sex in injury nature and body part are less obvious (Figure S1). By sex differences in code distribution were not significant for primary source (p = 0.35), significant for event (p= 0.045), marginal for injury nature (p = 0.054) and highly for body part injured (p = 0.005, Supplementary Table S2).

**Figure 1.**
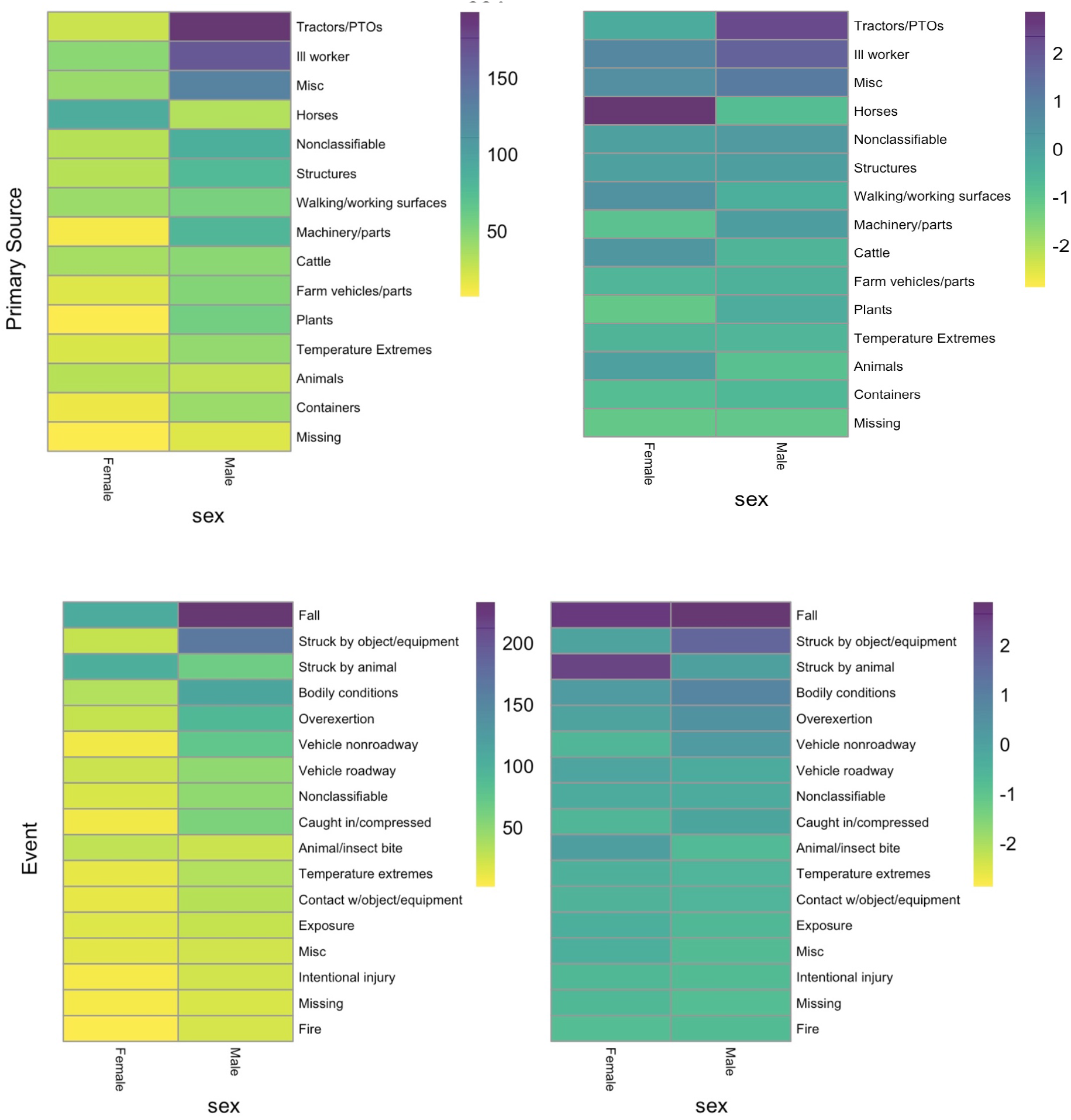
Heat maps illustrating differences in primary injury source (Source, top panels) and event (Event, bottom panels) by sex (Male, left; Female, right). Grouped codes are arranged by descending total frequency from left to right. Labels for grouped codes are shown along the bottom of each panel. Left column shows raw injury count maps to show highest counts; right shows counts scaled by sex to highlight primary injury source and event by sex.

### Injury by period

Injuries generally increased across the study period(s) from 2008-2011 to 2019-2022, with injury primary injury source (Source1) codes for ‘tractors/PTO’s’ and ‘Ill worker,’ the largest categories overall, peaking (purple) in the second period (2012-16) and then declining in the third (2019-2022) (Table 1). Given distributions in injuries by sex highlighted above, tractor injuries, primarily male, peaked during the second period (2012-2016), while large animal injuries, primarily female, show a relative maximum in the second period for cattle and the third period for horses. There was a notable increase in ‘unclassifiable’ sources in the third period (Figure 2, top panels). The injury event (Event) visualization shows a similar increase (green to blue to purple) across periods, with ‘Fall’ events being dominant across all periods and increasing sharply across the study period, especially for 2019-2022. Events associated with Temperature extremes increased slightly across periods (Figure 1, bottom panels). Injury Nature codes showed strong increases across periods for ‘pain’ and ‘traumatic injuries’ (Supplemental Figure S2); and injuries involving multiple body parts, or described simply as ‘body’ increased strongly across the study period (Supplemental Figure S2). By period differences in code distribution were highly significant for all OIICS coding levels (p< 0.001). In addition, injury count differences by period and sex, and period and age-category, were also highly significant (p<0.001; Table 1).

**Table 1.**
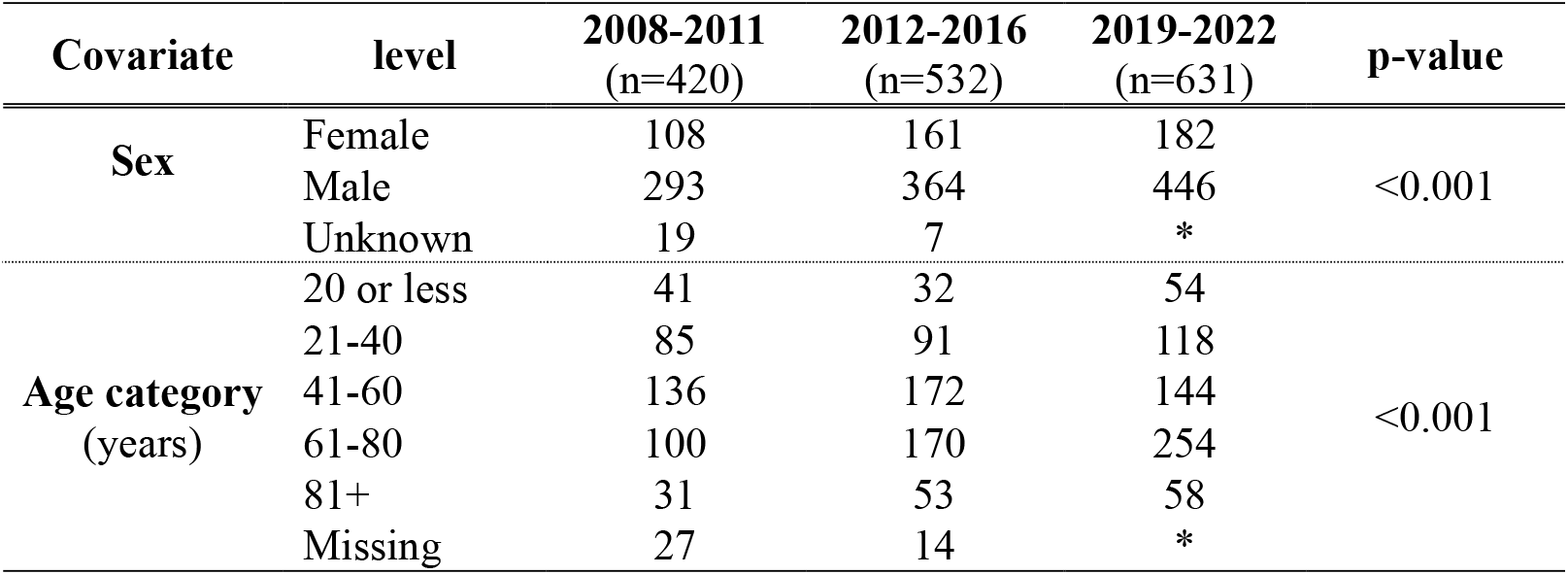
Injury counts across sex and age category, stratified by study sub-period; p-values from Chi-squared tests. Cell sizes less than 5 counts are redacted (*). Sample size: N=1,583.

| **Covariate** | **level** | **2008-2011**<br>**(n=420)** | **2012-2016**<br>**(n=532)** | **2019-2022**<br>**(n=631)** | **p-value** |
| --- | --- | --- | --- | --- | --- |
| **Sex** | Female | 108 | 161 | 182 | <0.001 |
|  | Male | 293 | 364 | 446 |  |
|  | Unknown | 19 | 7 | * |  |
| **Age category**<br>**(years)** | 20 or less | 41 | 32 | 54 | <0.001 |
|  | 21-40 | 85 | 91 | 118 |  |
|  | 41-60 | 136 | 172 | 144 |  |
|  | 61-80 | 100 | 170 | 254 |  |
|  | 81+ | 31 | 53 | 58 |  |
|  | Missing | 27 | 14 | * |  |

**Figure 2.**
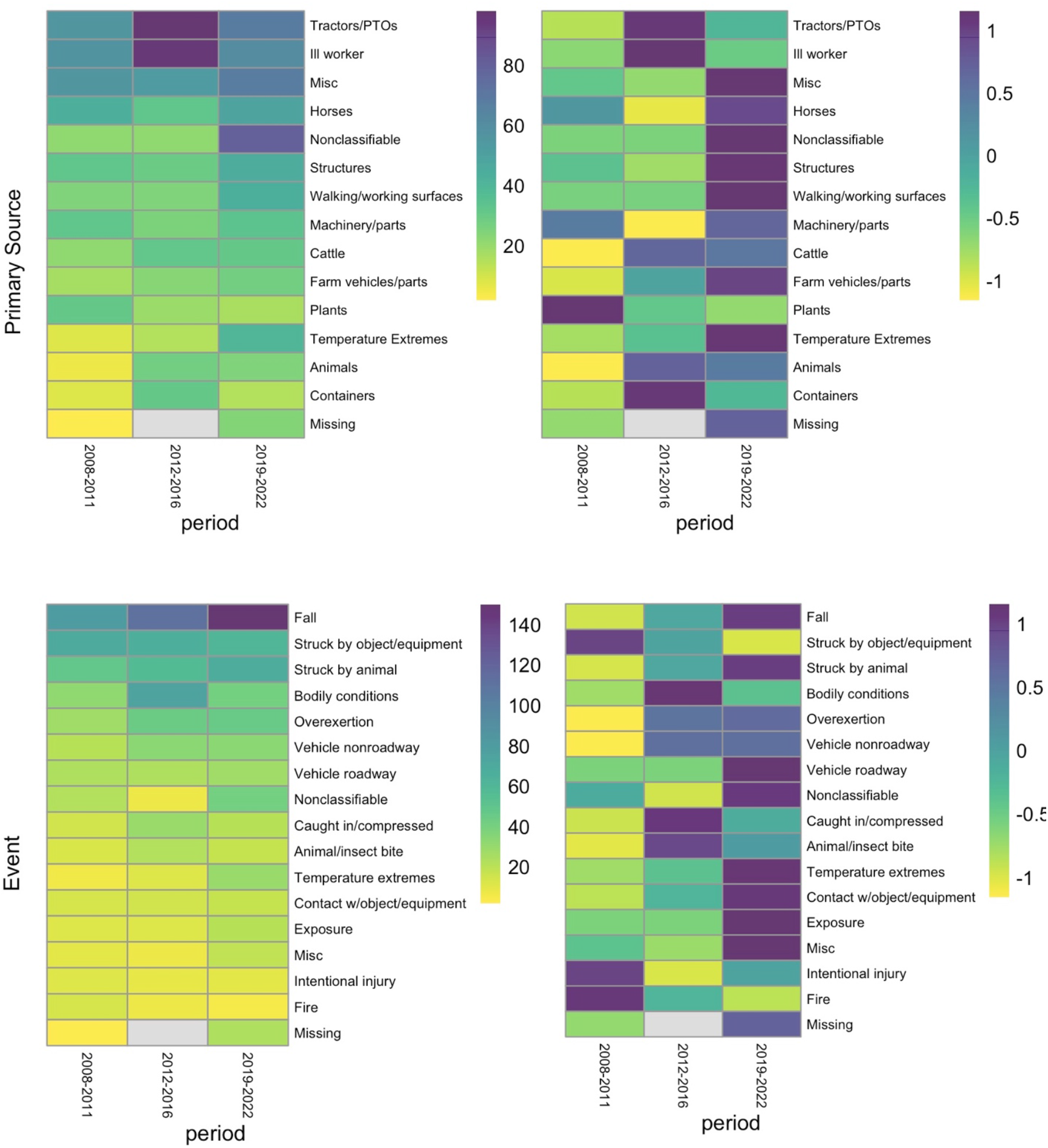
Heatmaps for OIICS primary Injury Source (source1, top panels) and Event (bottom panels) by **period of interest**; from left by column within each panel: 2008-2011, 2012-2016, and 2019-2022. Labels for grouped codes are shown on the left side of each panel and are arranged in descending order of appearance. Left panels are maps of raw counts, and right panels are scaled by code grouping to highlight which periods had the highest counts.

### Injury seasonality

We observed strong seasonality in most but not all codes, increasing from spring to a summer peak, then declining in the fall to a winter minimum (Figure 3). Again sources (Source1) involving tractors/PTO’s and ‘ill worker’ had the largest summer peaks. The scaled source map suggests walking/working surfaces were also problematic in the winter, perhaps due to falls on icy surfaces. Temperature extremes also show a winter peak under primary source (Figure 3, see top right panel). Injury events (Event) involving falls peaked in the summer but were dominant in count across all seasons, and events involving equipment, animals, overexertion, vehicles, and extreme temperatures also showed summer maxima (Figure 3, see bottom right panel). Injuries involving multiple body parts, head injuries, injuries to the trunk and upper extremities had summer peaks (Part). Injuries to the legs were more likely in the spring and fall seasons; foot and lower extremities in the spring and summer (Supplemental Figure S3). Note that these are not adjusted for population at risk, so are injury frequencies or counts. However, injury rates computed in a prior paper closely tracked frequencies (Jones, Scott et al. 2025). By season differences in code distribution were highly significant for primary injury source (p=0.007) and injury event (p=0.007); significant for body part injured (p = 0.039) and marginal for injury nature (p=0.07).

**Figure 3.**
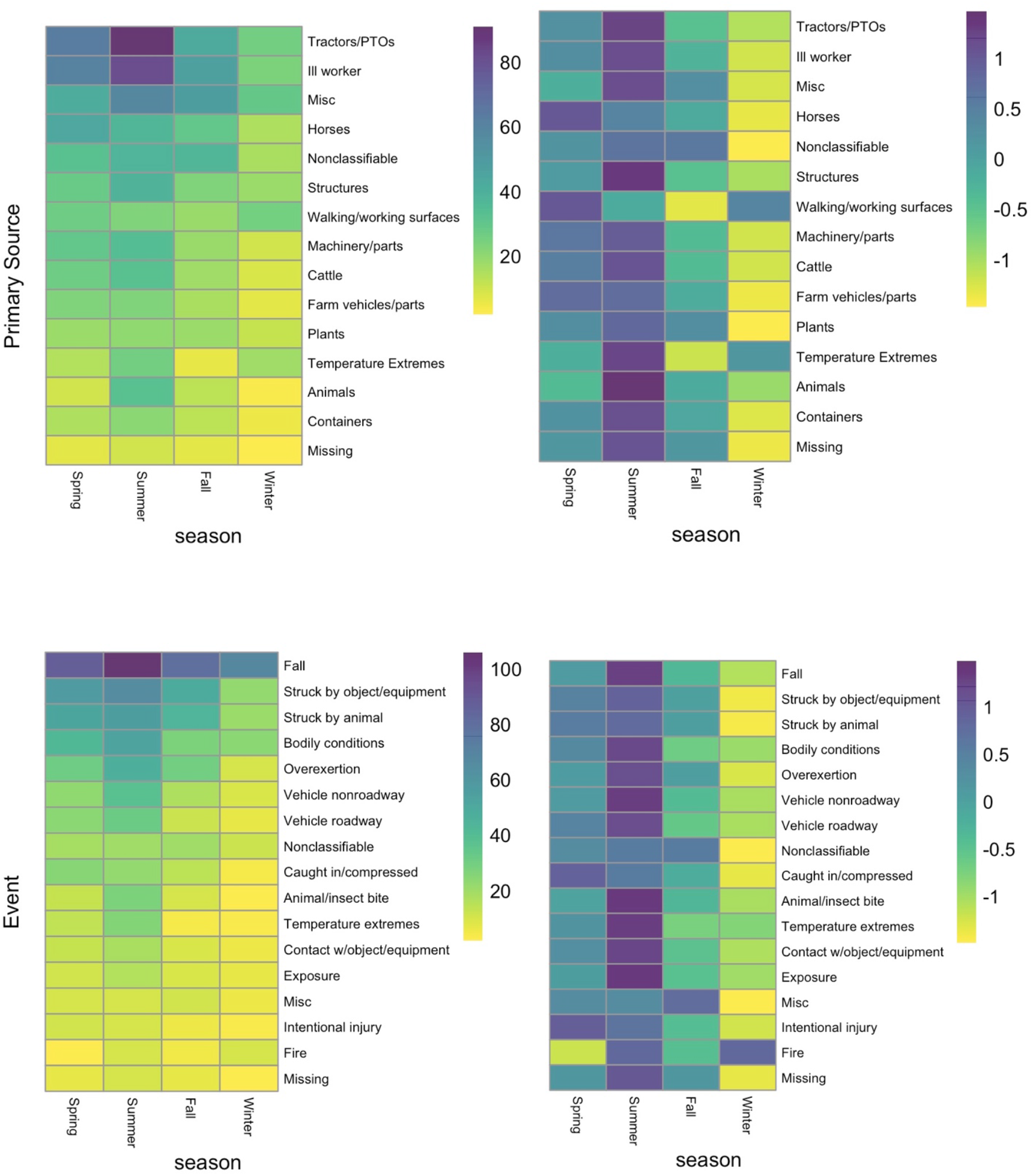
Heatmaps for OIICS primary Injury Source (source1, top panels) and Event (bottom panels) by **season**, from left in each panel: spring, summer, fall and winter. Labels for grouped codes are shown on the left side of each panel and are arranged in descending order by count occurrence. Left column shows raw count maps, and right column shows maps scaled by grouped label to highlight highest hits by season for each item. Listings of OIICS codes by label for each code level (Source, Event, Nature and Part) appear in Supplemental Table 1.

### Injury by age category

For the injury by age category, we show both unscaled (counts per cell) heat maps and mean-scaled heat maps (Figure 4; Supplementary Figures S4-S5). While unscaled maps show allow comparisons across all grouped codes and age categories; scaled maps, where cell-counts for each grouped code (row) are mean-standardized, facilitate comparison by age category within a particular grouped code. All maps show grouped codes in descending order by cell counts. In all code groupings, the young (ages 20 and under) are least often injured, however, this may not translate into dramatically lower injury rates, since we do not have age-specific population at risk numbers (Table 1). Likewise, if older workers represent a modest fraction of the agricultural working population, as they are over-represented among the injured, injury rates would likely be unusually high for this cohort. Oddly, the 41 to 60 year cohort showed increases from 2008 to a peak in 2016, followed by a subsequent decline in injuries.

**Figure 4.**
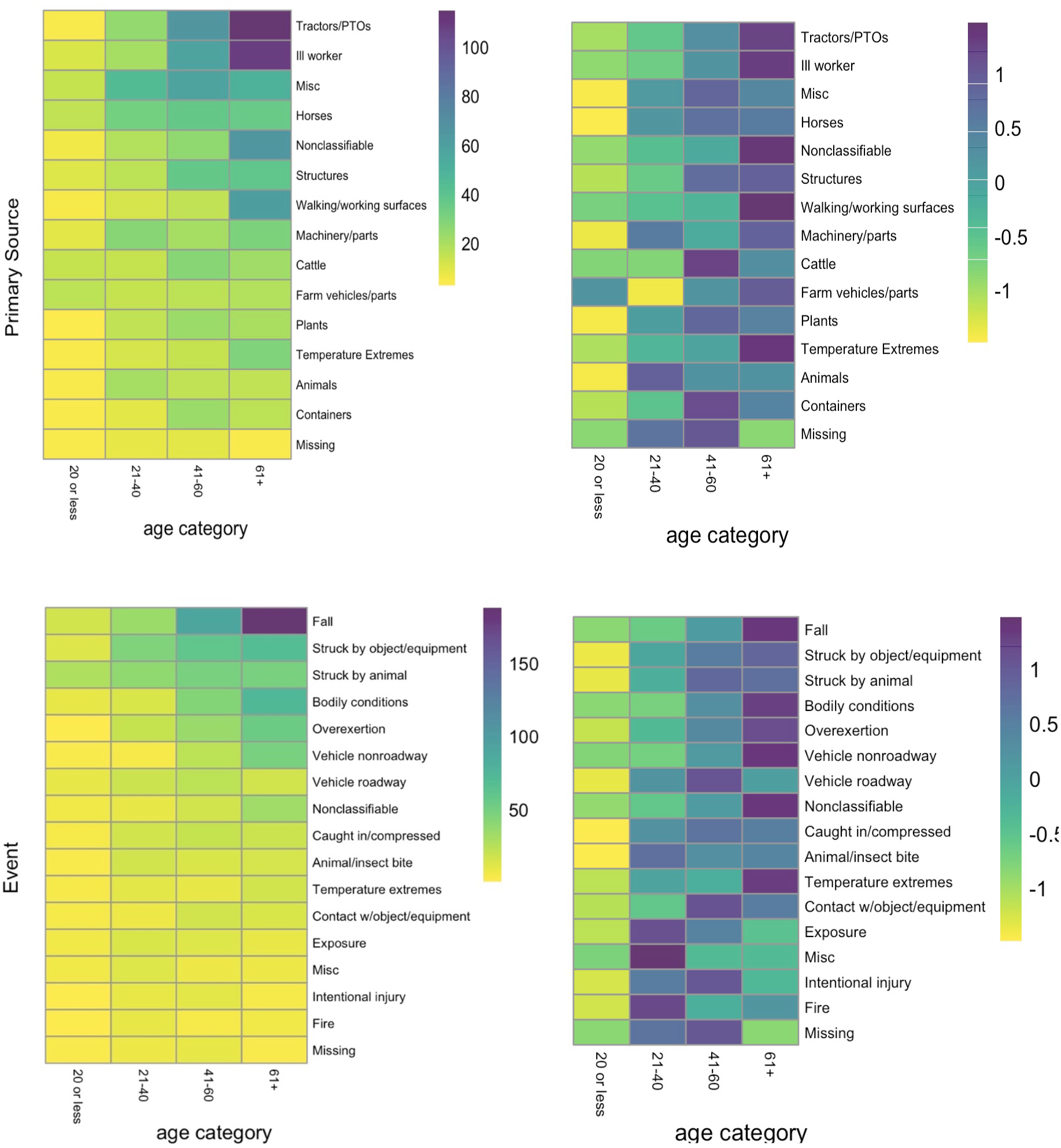
Heatmaps by **age category**. Primary Source (Source1) top row; Event, bottom row. Source and Event labels read across rows, and age categories read down columns. Age categories for ages 61 years and above are shown as one combined age category, “61+” due to relatively small number of injuries (see Table 1). Left column shows heat maps of injury counts falling into each grouped label/age category bin; right column is scaled across age categories to show, for each code grouping, relative ‘weights’ in each age category.

Maps showing counts again demonstrate that for primary source (Source), older workers (ages 61+ and 41 to 60 years) are more often injured by tractors/PTOs, or be reported as ‘ill worker’, to be injured on walking or working surfaces, or to report a Nonclassifiable source (Figure 4A). The scaled primary source plot suggests further nuance, that among workers whose source of injury is ‘machinery/parts,’ both the oldest cohort (61+) and the ages 21 to 40 cohort show higher weightings. Injuries by horses are almost equally spread between three cohorts older than 20 years; while injuries by cattle are more heavily weighted for the ages 41 to 60 cohort, and injuries by animals are more often experienced by the ages 21 to 40 cohort (Figure 4, top panels). Finally, ‘temperature extremes’ is most heavily weighted among the oldest cohort (61+).

The unscaled map for injury event (Event) confirms that, by far, a fall is the most common injury event and that falls are strongly associated with age (Figure 4, bottom panels). Scaled maps show that the oldest workers (61+) are also more likely to suffer ‘bodily conditions,’ overexertion, vehicle non-roadway, and non-classifiable events, as well as events associated with temperature extremes (Figure 4, right column). Meanwhile, the middle-aged cohort (ages 41 to 60) is the most likely to suffer vehicle roadway, contact with object/equipment, and intentional injury events, as well as to have missing event data. Ages 21 to 40 cohort is most likely to show fire or ‘exposure’ as the injury event.

For injury nature (Nature), the most dominant code grouping, “pain” increases dramatically with age on the count maps (Supplemental Figure S4, top). The scaled maps show a more nuanced picture, with all labels (aside from ‘Open wounds,’ ‘Poisoning’, and ‘Missing’) showing the heaviest “weights” (purple and blue) for the oldest cohort of workers (ages 61+, Supplemental Figure S4, bottom panels). Of note, the ages 21 to 40 cohort are most likely to show nature of injury as ‘poisoning,’ a code grouping that includes drug overdoses. The oldest cohort is most likely to have an injury of ‘non-classifiable’ nature, but least likely to have a missing code.

Body part injured is dominated by multi-injuries (injuries to multiple parts of the body, or merely ‘body’) and again it is primarily the older cohort (Supplemental Figure S5). Other common body parts injured include head, trunk, upper extremities, legs, and chest. The scaled map confirms this and also shows high rates of non-classifiable body part determinations and low missing codes for older workers. Missing units appear highest for the intermediate cohorts (ages 21 to 60). By age category differences in code distribution were highly significant for all OIICS coding levels (p< 0.001, Supplemental Table S3).

## Discussion

### Summary

We employed a novel time series dataset comprising 15 years of agricultural injuries (2008-2022, with a 2-year hiatus) mined from pre-care records (PCRs) from the state of Maine, then classified in detail using the OIICS classification system. A prior time series study showed a strong annual seasonality in injuries by month across all periods and demonstrated increasing seasonality-adjusted moving average injury rates across study periods (Jones, Scott et al. 2025b, submitted). We observe the same dynamics in the injury code groupings, though we report counts instead of rates since we do not have granular information on the populations at risk by sex or age category. With few exceptions, including primary source tractors/PTOs and ‘ill worker’, which peaked in the second period (2012-2016), most injury code levels increased across the three sub-periods considered and show strong seasonality, with maxima in summer and minima in winter. Male subjects had higher injury counts than females (Table 1) and tended to be injured by equipment, while females were more likely to be injured during interactions with large animals. Injuries increased with age category except for the very oldest cohort (age 81+, Table 1). ‘Falls’ increased across age categories from young workers to older workers, and older workers were most likely to suffer a fall irrespective of season.

Given Maine’s growing season it is unsurprising that we observe an increase in injuries during the summer months. Cold, snowy winters are more conducive to machinery repair and tending to animals, but crop production significantly increases in the summer and autumn. The agricultural landscape in Maine is dominated by small diversified farms, growing nearly the nation’s entire supply of wild blueberries, along with significant amounts of potatoes, broccoli, cauliflower, apples, and notable diary and livestock production.^28,29^ The observed increase in injuries in our data is consistent with other documented literature, and with increases in the risk exposure for agricultural work during the northeast growing season.^30,31^

Similarly, the observed ages and sex stratification are typical for the industry and type of work. Young workers and children can be at risk for agricultural injury given that the farm is often the homestead as well. Children routinely take on work-related tasks at a young age, and the complexity of such tasks can increase as children mature physically and mentally.^32,33^ Likewise, the concept of a normal “retirement age” is not always reasonable in agriculture. In fact, the latest Census of Agriculture (2022) showed the average age of Maine farmers has crept up to 57.5.^28^ The entire agricultural workforce is aging, and working well into someone’s 70’s and 80’s is routine.^34^ Agricultural tasks remain relatively segregated by sex. Men traditionally work more frequently with large equipment and it is common for women to take a greater role in animal care.^35^

With animal care, horse-associated injuries are particularly challenging for an occupational injury surveillance system, as it is difficult to discern work-related from recreational horse injuries.^36-39^ However as injury prevention recommendations are similar regardless of work-relatedness, we thought this injury source was important to capture and present here. Similar issues arise for other injury sources such as ATVs and small tractors, where it’s hard to discern use on a working farm from use for personal property maintenance.^40^ Increasing pain with age may be connected with the aging workforce, and the aging workforce trend is likely to continue. Researchers and those working in agriculture need to consider the special needs of this important demographic. Lastly, more than many other injury surveillance systems, PCRs provide injury scenario-related details that are critical to targeted action by injury surveillance specialists. PCRs should continue to be a data source for agricultural injury.

### Strengths and limitations

Our PCR-based dataset includes only injuries where EMS was activated, thus it may underestimate true injury counts. Less severe incidents that did not prompt emergency calls were not captured. In addition, incidents among agricultural workers with precarious immigration status, where seeking medical attention may jeopardize work status, are likely also omitted.

While the vast majority of PCR records have detailed narrative free-text accounts of an injury or illness scenario, it is also possible that agricultural injury records exist in the dataset that were not captured by the machine learning algorithm. Exclusion of coded records from 2017 (not available) and 2018 (omitted due to problems with duplications) omits a critical period that may explain the large rise in injury events between 2016 and 2019. Our novel dataset shows significant difference in injury scenarios and types by age category and sex, presented as frequencies. However, we cannot compute rates because we lack granular information about farm employment by sex or age category.

## Conclusions

We found that injury counts rose across each successive study period, and that injury dynamics were strongly seasonal, with a peak in the summer and a winter minimum. As with non-farming populations, falls are a prominent injury event that increase in frequency with age. Falls increased in frequency across the study period. Event and Nature subcodes differed significantly by sex, with males more likely to be injured by equipment and females by large animals, while primary injury Source and Body Part codes also varied significantly by age. This suggests that injury risks are not uniform across worker demographics, and tailored safety interventions by sex and age group may be more effective in reducing agricultural injuries.

## Data Availability

The data analyzed here are available in raw form from the Maine Bureau of Emergency Medical Services and are used under license for the current study. Those interested in these records may contact the Maine EMS Bureau. The workflow and code developed by the authors is available by a written request to the corresponding author.

https://www.maine.gov/ems/mefirs

## Author contributions

LEJ: conceptualization, data curation and cleaning, analytical plan, visualization, analysis, writing (original draft), manuscript development and revision; MK: visualization, writing (original draft), manuscript development and revision; NK: data selection, curation and cleaning; CH-R: project management, manuscript development and revision; ES: funding, project management, manuscript development and revision; and PJ: manuscript development and revision. All authors read and approved the final manuscript.

## Funding

Funding for this study was provided by the Centers for Disease Control and Prevention, National Institute for Occupational Safety and Health (CDC-NIOSH) Grant No. 2U54OH007542.

## Declarations

### Ethics approval and Consent to Participate

This study was approved by the Institutional Review Board of the Mary Imogene Bassett Hospital (Bassett Medical Center) and was conducted in accordance with the ethical standards of the Declaration of Helsinki. A waiver of consent to participate was approved by the IRB as this is a retrospective study of data extracted from text records licensed by the state of Maine EMS Bureau.

### Consent for Publication

Not applicable.

### Competing interests

The authors declare that they have no competing interests.

## Notes

### Competing Interest Statement

The authors have declared no competing interest.

