## Supplemental Information for "OIICS coding of agricultural injuries mined from Maine PCR records (2008-2022) reveals significant differences in injury source, event, and nature by age group and sex"

### Supplementary Information

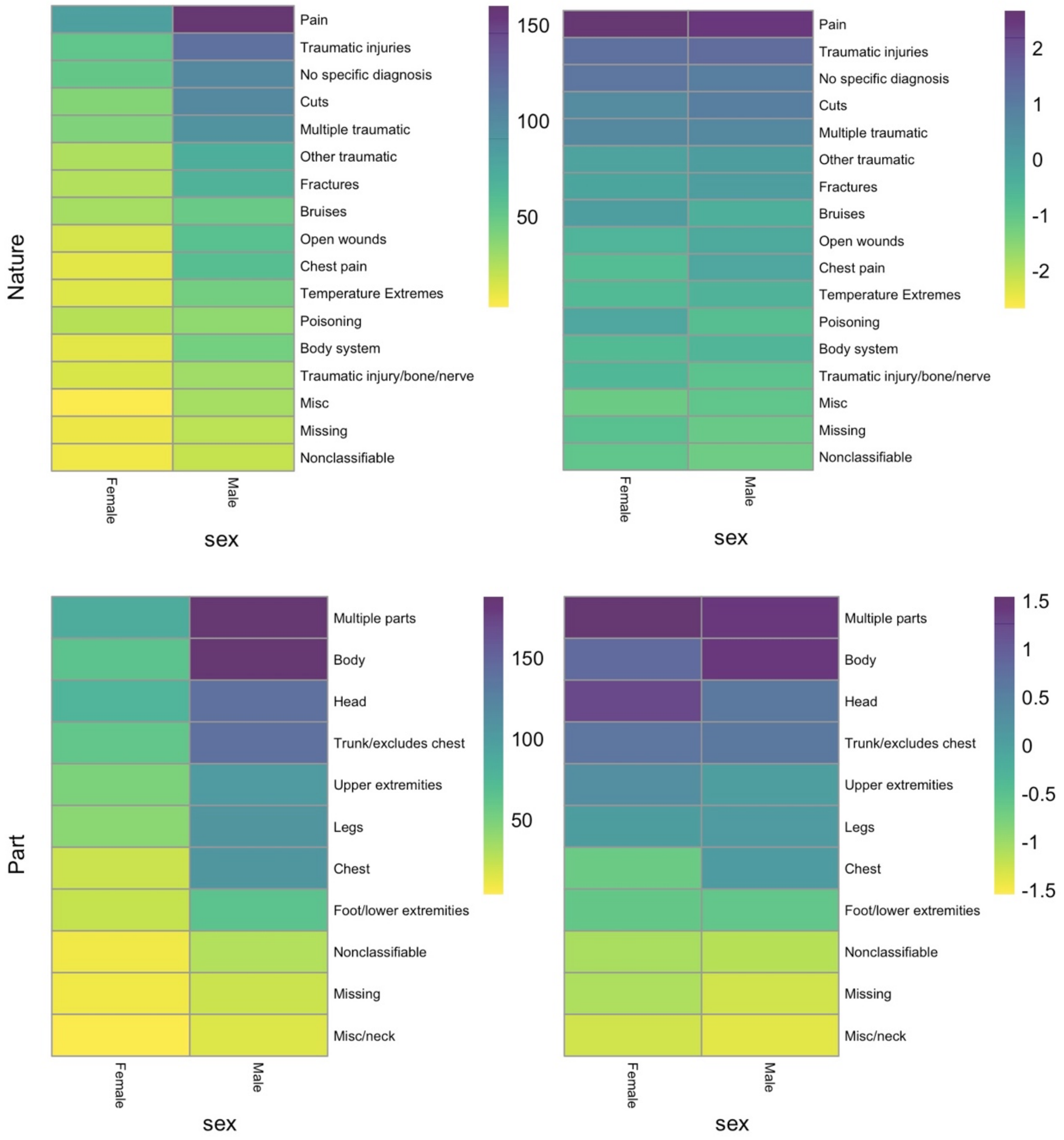


**Figure S1.** Heat maps illustrating differences in injury nature (Nature, top panel) and body part injured (Part, bottom panel) by sex (Male, left; Female, right). Grouped codes are arranged by descending total frequency from left to right. Labels for grouped codes are shown along the bottom of each panel. Right panels show count maps and left panels are scaled by sex to show which items occur more frequently for each sex.

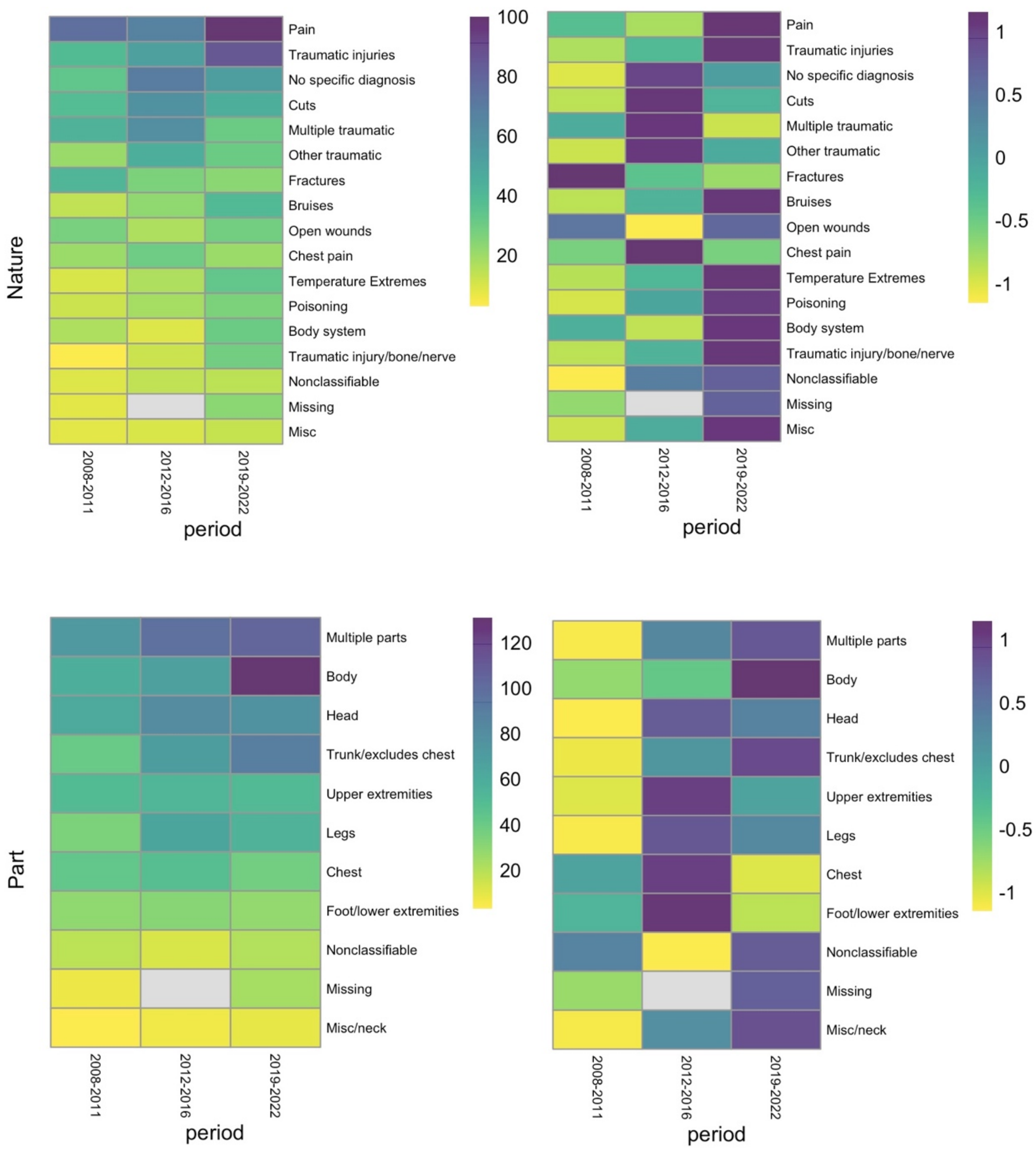


**Figure S2.** Heat maps for OIICS Injury Nature (Nature, top panels) and Body part injured (Part, bottom panels) by period of interest; from left by column within each panel: 2008-2011, 2012-2016, and 2019-2022. Labels for grouped codes are shown on the left side of each panel and are arranged in descending order by count frequency. Left hand panels are count maps to show global peaks in injury nature and body part by period. Right panels are scaled by grouped label to reveal peaks in each item by period. For a listing of OIICS codes by label for each code level (Source, Event, Nature and Part), please see Supplemental Table 1.

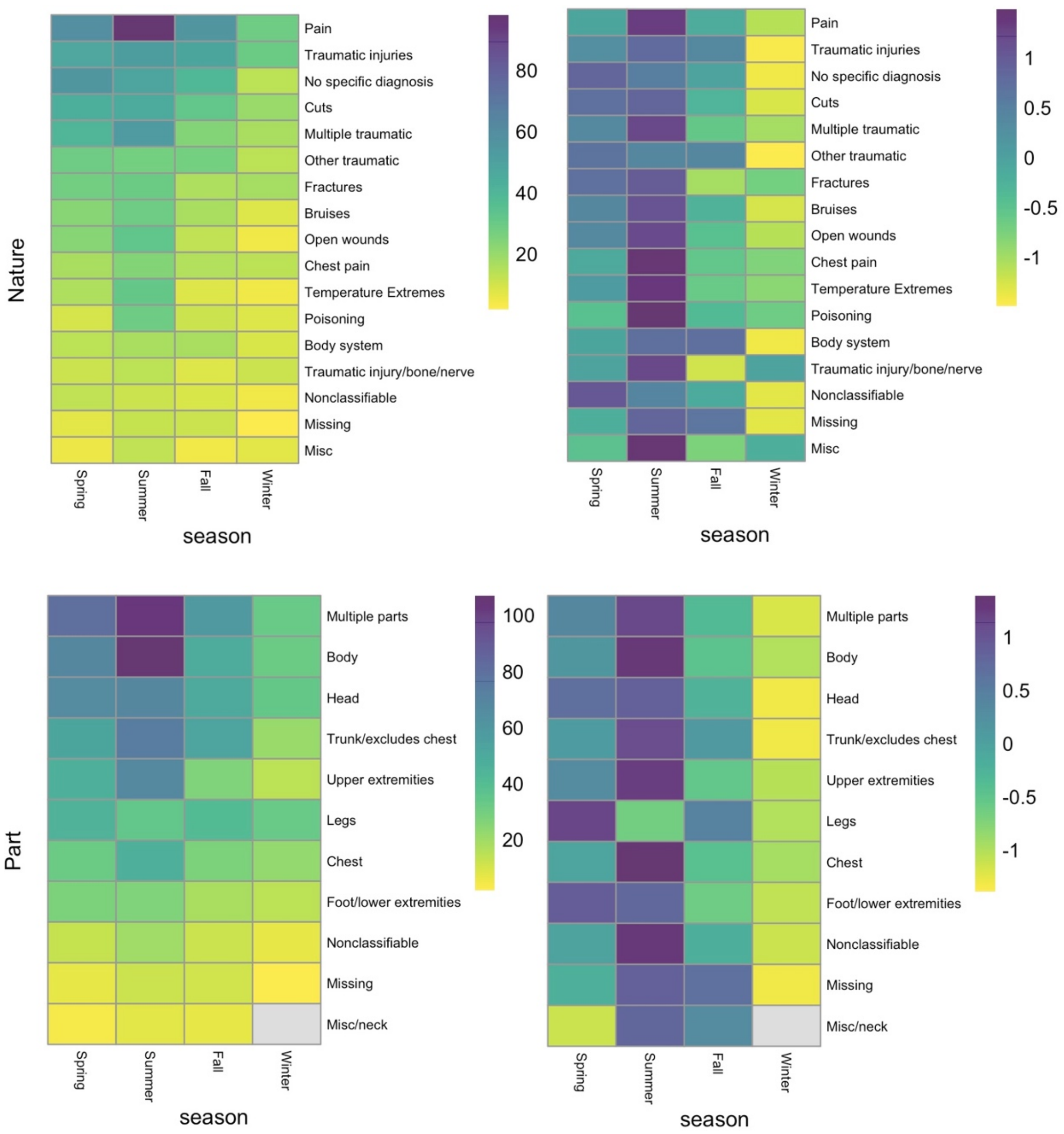


**Figure S3.** Heat maps for OIICS Injury Nature (Nature, top panels) and Body part injured (Part, bottom panels) by season; from left by column within each panel. Labels for grouped codes are shown on the left side of each panel and are arranged in descending order by count frequency. Left hand panels are count maps to show global peaks in injury nature and body part by season across all periods. Right panels are scaled by grouped label to reveal peaks in each item by season. For a listing of OIICS codes by label for each code level (Source, Event, Nature and Part), please see Supplemental Table 1.

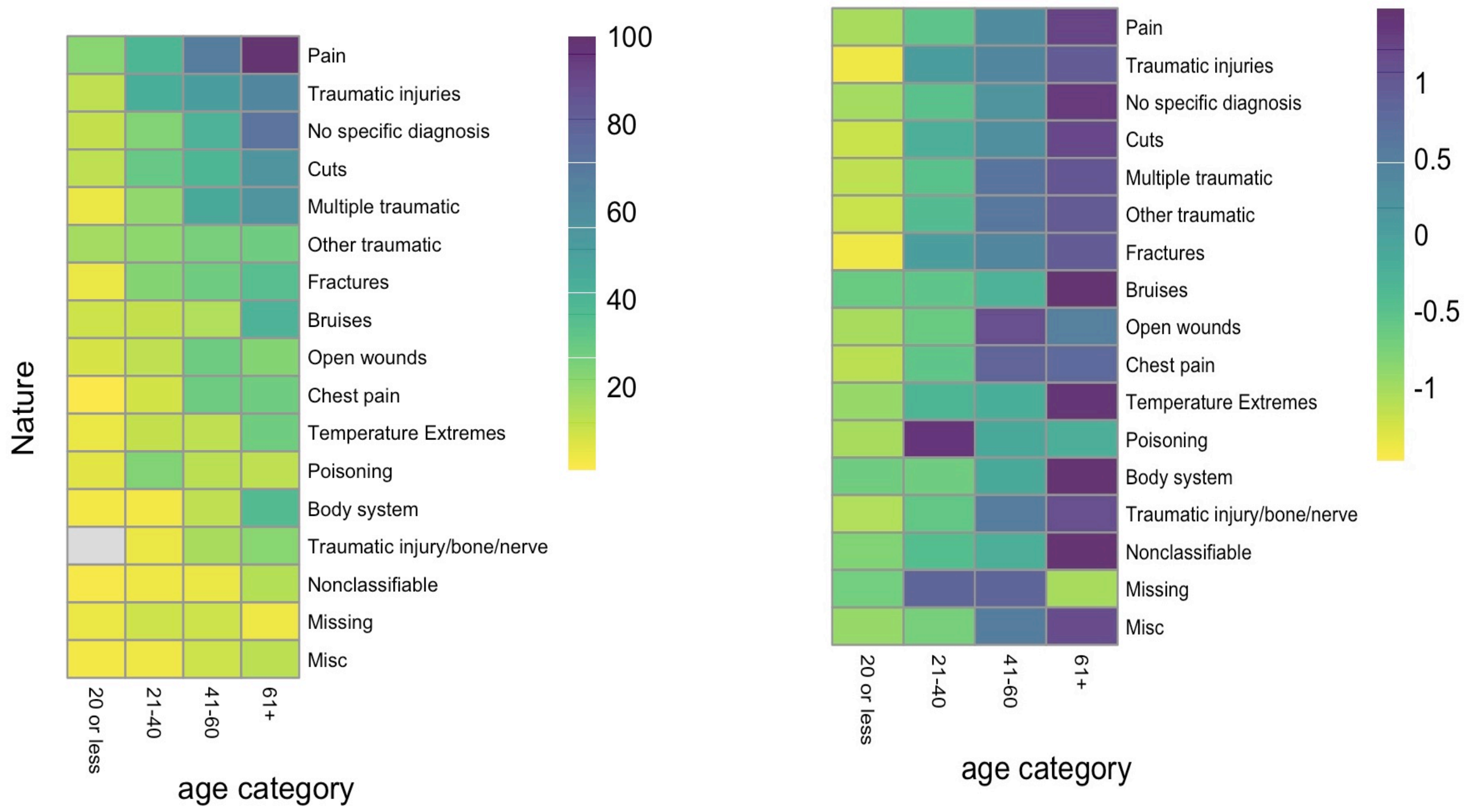


**Figure S4.** Heatmaps by age category, nature of injury (Nature). **Left,** heatmap of counts per cell; **Right,** scaled map (scaled along rows). Grouped labels are read across rows, and age categories read down columns. Age categories for ages 61 years and above are shown as one combined age category, "61+" due to relatively small number of injuries for ages 81 and above (see Table 1).

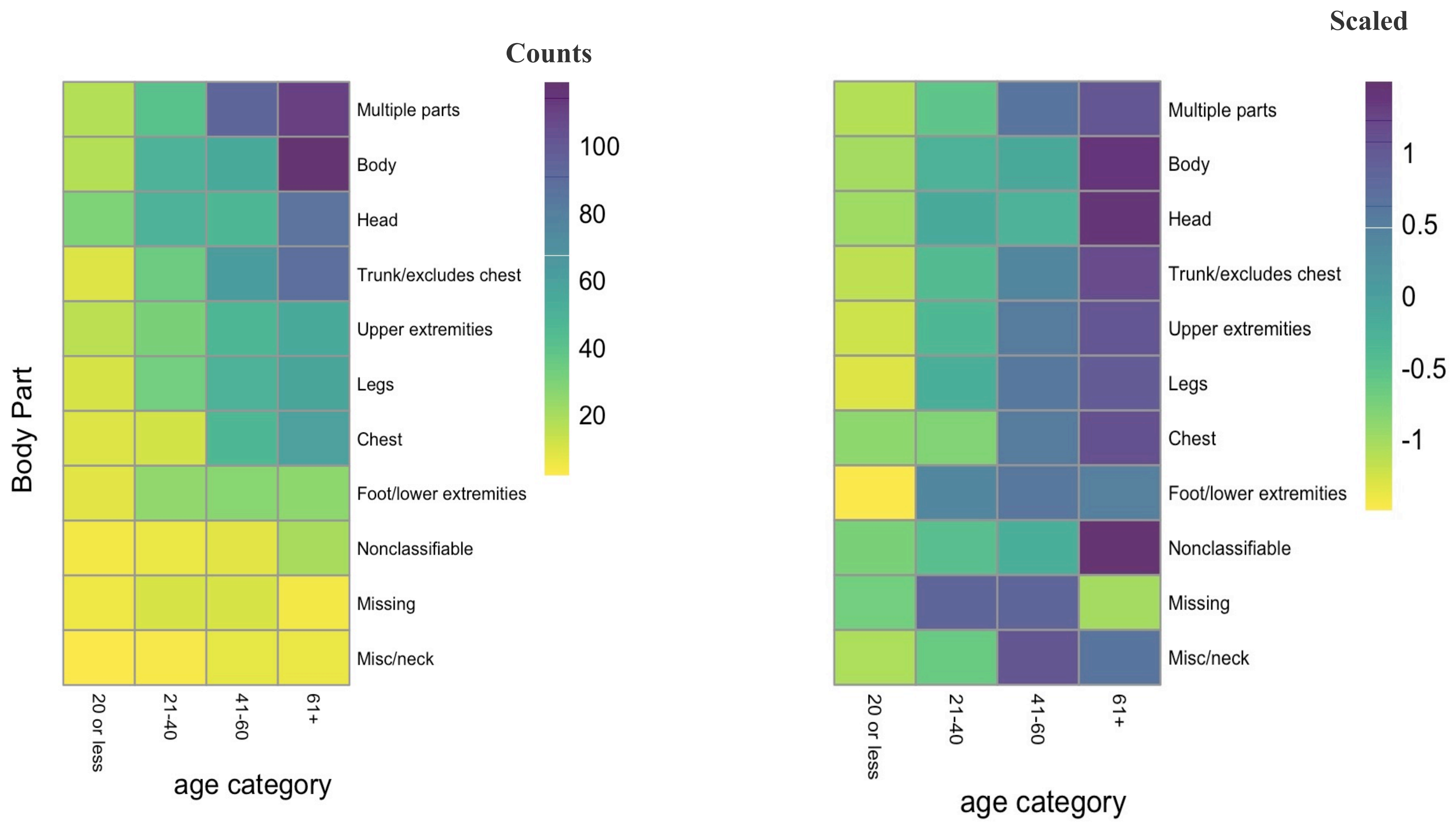


**Figure S5.** Heat maps of body part injured by age category. **Left,** heatmap of counts per cell; **Right,** scaled map (scaled along rows). Grouped labels are read across rows, and age categories read down columns. Age categories for ages 61 years and above are shown as one combined age category, "61+" due to relatively small number of injuries for ages 81 and above (see Table 1).

**Table S1.** Thematic OIICS codes groupings

| Structure | Thematic group | OIICS codes |
|---|---|---|
| Nature | Traumatic injury/bone/nerve | 11, 110, 118, 119 |
| | Traumatic injuries | 1, 12, 120, 121, 1211, 1212, 1219, 1231, 1232, 124, 128, 129, 150, 151, 1510, 1520, 1521, 1522, 1523, 1529, 16, 160, 161, 162, 168, 1681, 1689, 10 |
| | Fractures | 111 |
| | Open wounds | 13, 130, 138, 139, 131, 1311, 1312, 1319, 133, 134 |
| | Cuts | 132 |
| | Bruises | 140, 141, 143, 148, 149 |
| | Temperature extremes | 1710, 1712, 172, 1720, 1721, 1722, 1723, 1725, 1728, 1729 |
| | Multiple traumatic (injury) | 18, 180, 181, 183, 1830, 1832, 1839, 185, 189, 183, 1830, 1832, 1839, 1840, 1841, 1849 |
| | Other traumatic (injuries)[a] | 190, 191, 192, 193, 194, 1950, 197, 1971, 1973, 1978, 1979, 199, 1991, 1999 |
| | Poisoning | 196, 1960, 1961, 1962, 1963, 1964, 1965, 1966, 1967, 1968, 1969 |
| | Pain | 1972 |
| | Body system (diseases) | 20, 2912, 2919, 299, 2231, 2259, 232, 233, 2330, 2331, 2332, 2350, 2352, 2353, 2359, 2361, 2362, 237, 2379, 2389, 240, 2443, 2449, 252, 253, 2721, 2722, 2723, 279 |
| | No specific diagnosis (, signs and symptoms) | 50, 510, 5121, 5123, 5141, 5149, 5150, 5151, 5158, 5159, 5171, 5174, 5179, 518, 525, 58, 511, 5110, 5111, 5112, 5113, 5114, 5115, 5116, 5118, 5119 |
| | Chest pain | 5164 |
| | Misc[b] | 3129, 516, 5160, 5161, 5168, 5169, 60, 7, 8, 62, 620, 6210, 6213 |
| | Nonclassifiable | 9999 |
| Body Part | Head | 1, 10, 11, 110, 111, 112, 113, 118, 119, 12, 13, 130, 131, 132, 1330, 1333, 1339, 134, 135, 1361, 1368, 138, 18, 19 |
| | Trunk/excludes chest | 3, 30, 32, 320, 321, 322, 328, 329, 33, 330, 331, 332, 333, 3350, 339, 341, 342, 343, 344, 348, 349, 38, 39 |
| | Chest | 31, 310, 311, 312, 313, 315, 318, 319 |
| | Upper extremities | 4, 40, 41, 420, 421, 422, 423, 428, 4281, 4289, 43, 44, 440, 442, 4420, 4422, 4429, 448, 449, 481, 4819, 4839, 484 |
| | Foot/lower extremities | 50, 52, 53, 530, 5310, 5311, 5319, 532, 538, 5810, 5819, 582, 5830, 5839, 589, 59 |
| | Legs | 51, 510, 511, 512, 513, 518, 5180, 5181, 5189 |
| | Multiple parts | 8, 80, 81, 82, 83, 84, 85, 86, 87, 89, 891, 899 |
| | Body | 6 |
| | Misc | 2, 20, 21, 29, 914 |
| | Nonclassifiable | 9999 |
| Event - exposure | Intentional injury | 1110, 1111, 1112, 1113, 1114, 1119, 112, 1120, 1121, 1122, 1125, 1126, 1129 |
| | Animal/insect bite | 13, 130, 131, 1310, 1311, 1313, 1319, 1389, 139 |
| | Struck by animal | 132, 1320, 1321, 1322, 1323, 1324, 1329 |
| | Vehicle roadway | 20, 260, 2610, 2611, 2612, 2613, 2614, 2615, 2619, 2621, 2622, 2623, 2629, 2631, 2632, 2636, 2639 |
| | Vehicle non-roadway | 270, 2710, 2721, 2729, 273, 2730, 2731, 2732, 2734, 2736, 2737, 2738, 2739 |

| | | |
|---|---|---|
| | Fire | 310, 311, 312, 313, 314, 317 |
| | Fall | 40, 4420, 4421, 49, 43, 430, 4310, 4311, 4312, 4313, 4314, 4320, 4321, 4322, 4323, 4326, 433, 4330, 4331, 4332, 4333, 4334, 4335, 4336, 4337, 42, 420, 421, 4210, 4211, 4212, 4213, 4214, 4219, 422, 423, 424, 429 |
| | Exposure | 50, 53, 541, 57, 59, 510, 5110, 5111, 5112, 55, 550, 551, 552, 5520, 5521, 5522, 553, 554, 5540, 558, 559, 561, 563, 569 |
| | Temperature extremes | 531, 532 |
| | Contact w(ith)/object/equipment | 60, 630, 6310, 6312, 6320, 6321, 6322, 6329, 639, 655, 659, 663, 679, 69 |
| | Caught in/compressed | 64, 640, 641, 6410, 6411, 6412, 642, 643, 644, 649 |
| | Struck by object/equipment | 62, 620, 621, 6210, 6211, 6212, 6213, 6214, 6215, 6219, 6221, 6223, 6229, 6240, 6241, 6242, 6243, 6249, 625, 6250, 6252, 6259, 6260, 6261, 6269, 629, 621, 6210, 6211, 6212, 6213, 6214, 6215, 6219, 623, 6230, 6231, 6233, 6239 |
| | Overexertion | 70, 722, 723, 729, 78, 79, 71, 710, 711, 7110, 7111, 7112, 712, 7120, 7121, 7122, 713, 7130, 7132, 7141, 718, 719, 730, 7310, 7311, 7313, 7332, 735, 7351, 7360, 7361, 7370, 738, 7391, 7399 |
| | Bodily conditions | 74 |
| | Miscellaneous[c] | 2311, 2313, 2314, 2319, 24, 241, 2412, 2429, 244, 2440, 2442, 322, 4121, 4129, 10, 1214, 1220, 1222, 1224 |
| | Nonclassifiable | 9999 |
| Primary source | Animals (other than cattle/horses) | 510, 5112, 5130, 5131, 5136, 515, 5152, 5156, 5157, 5159, 520, 5212, 5216, 529 |
| | Cattle | 5153 |
| | Horses | 5154 |
| | Plants | 580, 582, 586, 587, 5870, 5871, 5872, 5873, 5879, 5891 |
| | Temperature extremes | 9261, 9273, 9262, 9292 |
| | Containers | 2111, 2112, 2114, 2115, 2116, 2117, 2118, 2122, 2131, 216, 2218 |
| | Ill worker | 560, 561, 562, 569 |
| | Machinery/parts | 30, 310, 3110, 3111, 3112, 3113, 3119, 3122, 3123, 3124, 313, 3130, 3131, 3133, 3199, 320, 3211, 3212, 3221, 3222, 3233, 3235, 3241, 3312, 3314, 3334, 3336, 339, 3421, 347, 3569, 3573, 3991, 3994, 440, 4417, 4418, 4422, 4423, 4429, 4432 |
| | Tractors/PTOs | 8630, 8631, 8633, 8634, 8639 |
| | Structures[d] | 60, 6171, 6220, 6221, 6222, 6229, 6251, 633, 634, 6340, 6342, 6350, 6352, 639, 6399, 6511, 6519, 652, 6520, 6522, 6523, 6524, 6529, 653, 654, 6540, 6543, 6544, 6550, 6551, 6562, 6569, 4110, 4120, 4123, 4130, 4132, 4135, 4139, 4150, 4151, 4152, 4153, 4198, 4199 |
| | Walking/working surfaces | 66, 660, 6610, 6611, 6612, 662, 6620, 6621, 6622, 6629, 663, 6630, 6631, 6639, 6650, 6671, 6676, 6692, 6699 |
| | Farm vehicles/parts | 8420, 8421, 8423, 8424, 8429, 8431, 8611, 8621, 8629, 873, 874, 879, 850, 852, 853, 859, 4813, 4819, 4821, 4829, 483, 484, 4850, 4852, 4853, 489 |
| | Misc[e] | 40, 4211, 4212, 4220, 4223, 4224, 4226, 2233, 1, 10, 1115, 139, 1469, 150, 151, 160, 1641, 1741, 1840, 1841, 1842, 1848, 19, 7831, 790, 924, 9240, 9241, 9243, 926, 9272, 9296, 9297, 781, 7811, 7819, 70, 710, 7124, 7125, 7127, 7131, 7133, 7151, 7159, 7192, 7193, 7194, 7199, 7211, 7221, 7260, 7261, 4432, 74, 740, 742, 7420, 7421, |

| | |
|---|---|
| | 7422, 7423, 552, 554, 556, 7529, 570, 5711, 5712, 5719, 5721, 576, 450, 451, 46, 460, 470, 9112, 940, 9414, 949, 9521, 80, 84, 840, 8410, 8413, 8416 |
| Nonclassifiable | 9999 |

*Notes:*

[a] Other traumatic injuries include suffocation, drownings, electrocution, …

[b] Misc is miscellaneous. Miscellaneous in nature includes zoonotic diseases, mental disorder, other/multiple diseases, respiratory/chest symptoms, and exposure to disease though not ill.

[c] Miscellaneous in events include animal powered vehicles and pedestrian-vehicle incidents, explosions, slips and trips, and unintentional injury by person.

[d] Structures refer to man-made structures, such as buildings, bridges,…, and their materials.

[e] Miscellaneous in primary source include chair, chemicals, defense devices and instruments, environment, fasteners and ropes, firearms, hand-tools and their parts, ladders, minerals, orthopedics, other human, materials or sources, and vehicles.

**Table S2.** OIICS subcode distributions by thematic label, stratified by sex (N = 1554 subjects; female = 451; male = 1103; omits the 29 subjects with sex = ‘unknown’ due to small cell sizes). Cell sizes less than 5 counts are redacted (*). P-values are from groupwise comparisons chi-square tests and are estimated via Monte Carlo (10,000 simulations). For a listing of OIICS codes assigned to each thematic label, please see Supplementary Table S1.

| Subcode | level | Female | Male | p-value |
|---|---|---|---|---|
| | Animals | 31 | 28 | |
| | Cattle | 37 | 46 | |
| | Containers | 14 | 40 | |
| | Farm vehicles/parts | 18 | 50 | |
| | Horses | 90 | 32 | |
| | Ill worker | 47 | 164 | |
| | Machinery/parts | 11 | 80 | |
| **Source1** | Miscellaneous | 42 | 131 | 0.35 |
| | Missing | 7 | 19 | |
| | Nonclassifiable | 31 | 88 | |
| | Plants | 7 | 58 | |
| | Structures | 31 | 77 | |
| | Temperature Extremes | 20 | 43 | |
| | Tractors/PTOs | 25 | 193 | |
| | Walking/working surfaces | 40 | 54 | |
| | Animal/insect | 28 | 24 | |
| | bodily conditions | 33 | 113 | |
| | Caught in/compressed | 8 | 58 | |
| | Contact w/object/equipment | 12 | 32 | |
| | Exposure | 16 | 27 | |
| | Fall | 106 | 233 | |
| | Fire | * | 21 | |
| | Intentional injury | 7 | 22 | |
| **Event** | Miscellaneous | 15 | 22 | *0.045* |
| | Missing | 7 | 19 | |
| | Nonclassifiable | 18 | 49 | |
| | Overexertion | 26 | 92 | |
| | Struck by animal | 101 | 67 | |
| | Struck by Object/Equipment | 26 | 164 | |
| | Temperature extremes | 13 | 33 | |
| | Vehicle non-roadway | 9 | 77 | |
| | Vehicle roadway | 24 | 50 | |

| Subcode | level | Female | Male | p-value |
|---|---|---|---|---|
| | Body system | **12** | **44** | |
| | Bruises | 28 | 49 | |
| | Chest pain | 11 | 59 | |
| | Cuts | 39 | 102 | |
| | Fractures | 24 | 68 | |
| | Miscellaneous | * | 28 | |
| | Missing | 9 | 22 | |
| | Multiple traumatic | 40 | 92 | |
| **Nature** | No specific diagnosis | 51 | 102 | *0.054* |
| | Nonclassifiable | 7 | 20 | |
| | Open wounds | 16 | 57 | |
| | Other traumatic | 26 | 70 | |
| | Pain | 82 | 160 | |
| | Poisoning | 23 | 35 | |
| | Temperature Extremes | 13 | 46 | |
| | Traumatic injuries | 53 | 119 | |
| | Traumatic injury/bone/nerve | 14 | 30 | |
| | Body | 65 | 188 | |
| | Chest | 22 | 106 | |
| | Foot/lower extremities | 23 | 65 | |
| | Head | 78 | 139 | |
| | Legs | 43 | 108 | |
| ***Part*** | Miscellaneous/neck | * | 15 | 0.005 |
| | Missing | 9 | 22 | |
| | Multiple parts | 86 | 188 | |
| | Nonclassifiable | 10 | 29 | |
| | Trunk/excludes chest | 61 | 140 | |
| | Upper extremities | 50 | 103 | |
| **N** | | **451** | **1103** | **(1554)** |

**Table S3.** OIICS subcode distributions by thematic label, stratified by Age Category (N = 1554 subjects). P-values are from groupwise comparisons chi-square tests and are estimated via Monte Carlo (10,000 simulations). Nonzero cell sizes less than 5 counts are redacted (*). Note: Age, and thus age category, include(s) 44 missing units, hence the discrepancies with other totals.

| OIICS subcode | Level | Age Category (years) | | | | p-value |
|---|---|---|---|---|---|---|
| | | 20 or less | 21-40 | 41-60 | 61+ | |
| Primary Source | Animals | * | 21 | 16 | 17 | <0.0001 |
| | Cattle | 15 | 15 | 28 | 23 | |
| | Containers | * | 9 | 23 | 18 | |
| | Farm vehicles/parts | 17 | 15 | 17 | 18 | |
| | Horses | 16 | 32 | 37 | 38 | |
| | Ill worker | 11 | 21 | 59 | 120 | |
| | Machinery/parts | 9 | 28 | 21 | 32 | |
| | Miscellaneous | 15 | 45 | 59 | 51 | |
| | Missing | * | 8 | 9 | 5 | |
| | Nonclassifiable | 6 | 18 | 26 | 68 | |
| | Plants | * | 16 | 23 | 22 | |
| | Structures | 10 | 17 | 37 | 43 | |
| | Temperature Extremes | * | 12 | 15 | 31 | |
| | Tractors/PTOs | * | 25 | 66 | 119 | |
| | Walking/working surfaces | 5 | 12 | 16 | 61 | |
| Injury Event | Animal/insect bite | * | 18 | 15 | 17 | <0.0001 |
| | Bodily conditions | 10 | 14 | 44 | 79 | |
| | Caught in/compressed | 5 | 17 | 20 | 23 | |
| | Contact w/object/equipment | 5 | 8 | 17 | 14 | |
| | Exposure | 7 | 14 | 12 | 9 | |
| | Fall | 18 | 35 | 90 | 193 | |
| | Fire | * | 9 | 5 | 7 | |
| | Intentional injury | * | 9 | 11 | 6 | |
| | Miscellaneous | 7 | 13 | 8 | 8 | |
| | Missing | * | 8 | 9 | 5 | |
| | Nonclassifiable | 6 | 10 | 18 | 34 | |
| | Overexertion | * | 20 | 36 | 61 | |
| | Struck by animal | 28 | 39 | 49 | 50 | |
| | Struck by object/equipment | 12 | 45 | 61 | 71 | |
| | Temperature extremes | 5 | 11 | 10 | 19 | |
| | Vehicle non-roadway | * | 5 | 23 | 50 | |
| | Vehicle roadway | 10 | 19 | 24 | 20 | |

| OICCS Subcode | Level | Age Category (years) | | | | p-value |
|---|---|---|---|---|---|---|
| | | **20 or less** | **21-40** | **41-60** | **61+** | |
| Injury Nature | Body system | * | * | 12 | 39 | |
| | Bruises | 10 | 11 | 15 | 41 | |
| | Chest pain | * | 9 | 29 | 31 | |
| | Cuts | 12 | 30 | 39 | 60 | |
| | Fractures | 5 | 23 | 28 | 36 | |
| | Miscellaneous | * | * | 10 | 13 | |
| | Missing | 5 | 10 | 10 | 6 | |
| | Multiple traumatic | 5 | 20 | 47 | 58 | |
| | No specific diagnosis | 11 | 24 | 42 | 78 | <0.001 |
| | Nonclassifiable | * | * | 5 | 14 | |
| | Open wounds | 8 | 12 | 29 | 24 | |
| | Other traumatic | 17 | 21 | 26 | 28 | |
| | Pain | 22 | 39 | 68 | 107 | |
| | Poisoning | 6 | 24 | 13 | 13 | |
| | Temperature Extremes | 5 | 11 | 12 | 30 | |
| | Traumatic injuries | 12 | 44 | 51 | 65 | |
| | Traumatic injury/bone/nerve | 0 | 5 | 16 | 23 | |
| Body Part Injured | Body | 17 | 49 | 55 | 130 | |
| | Chest | 9 | 11 | 46 | 62 | |
| | Foot/lower extremities | 8 | 25 | 27 | 28 | |
| | Head | 30 | 50 | 46 | 90 | |
| | Legs | 10 | 33 | 50 | 59 | |
| | Miscellaneous/neck | * | * | 7 | 6 | 0.001 |
| | Missing | 5 | 10 | 10 | 6 | |
| | Multiple parts | 17 | 41 | 93 | 116 | |
| | Nonclassifiable | * | 6 | 8 | 20 | |
| | Trunk/excludes chest | 9 | 35 | 63 | 90 | |
| | Upper extremities | 16 | 31 | 47 | 59 | |
| **N** | | **127** | **294** | **452** | **666** | **(1539)** |
